# Chronic lesion expansion persists in secondary progressive multiple sclerosis and is associated with neurodegeneration

**DOI:** 10.64898/2026.09.23.26363839

**Authors:** Samuel Klistorner, Michael H Barnett, Alexander Klistorner

**Affiliations:** Brain and Mind Centre, University of Sydney, Sydney, New South Wales, Australia; Sydney Neuroimaging Analysis Centre, Camperdown, New South Wales, Australia; Save Sight Institute, Sydney Medical School, University of Sydney, Sydney, New South Wales, Australia

## Abstract

**Background:** Secondary progressive multiple sclerosis (SPMS) is traditionally viewed as predominantly neurodegenerative, with limited inflammatory activity. However, chronic active lesions characterised by persistent rim inflammation and gradual expansion may continue to contribute to tissue injury.

**Objectives:** To characterise chronic lesion dynamics in SPMS and determine whether chronic lesion expansion contributes to tissue damage and neurodegeneration. Thirty-one patients with SPMS underwent longitudinal MRI including T1-weighted, FLAIR and diffusion imaging. White matter lesions were manually segmented and chronic lesion tissue expansion (CLTE) was quantified as annualised percentage change. Associations between lesion expansion, mean diffusivity (MD) changes and brain atrophy were assessed using correlation, regression and mixed-effects modelling.

**Results:** CLTE remained prominent, with a mean annual expansion rate of 7.1 ± 4.1%. Expansion substantially exceeded the contribution of newly formed lesions and occurred at rates comparable to relapsing multiple sclerosis. CLTE demonstrated moderate intra-patient clustering (ICC = 0.23). MD increased significantly within lesions during follow-up and was associated with lesion expansion. Chronic lesion expansion was also strongly associated with ventricular enlargement (partial r = 0.72, p < 0.001).

**Conclusions:** Chronic lesion expansion persists in SPMS and represents a major source of tissue injury associated with microstructural degeneration and brain atrophy.

## Introduction

Secondary progressive multiple sclerosis (SPMS) is characterised by gradual and irreversible neurological deterioration that is only weakly associated with new inflammatory lesion formation. In this phase of disease, progression is thought to be driven predominantly by chronic, compartmentalised inflammatory and neurodegenerative processes within the central nervous system.[1][2][3] One of the principal pathological substrates of this process is chronically active or slowly expanding lesions, which are characterised by persistent microglial activation at their rim and progressive tissue destruction over time.[4][5][6]

In relapsing–remitting multiple sclerosis (RRMS), chronic inflammation at the rim of established lesions and corresponding chronic lesion tissue expansion (CLTE) are well established and have been linked to disability progression and tissue loss.[7][8] Imaging and neuropathological studies have demonstrated that a subset of chronic white matter lesions continues to expand over time, reflecting ongoing smouldering inflammatory activity.[9] However, whether this process persists to a similar extent in secondary progressive multiple sclerosis (SPMS) remains unclear, particularly as overt inflammatory activity declines in the progressive phase of the disease.

Most imaging studies in SPMS have focused on global measures such as brain atrophy, lesion burden and clinical progression, with limited direct assessment of chronic lesion dynamics.

As a result, the extent to which chronic lesion expansion contributes to ongoing tissue injury in established SPMS is not well characterised.[8]

Beyond the overall presence of chronic lesion activity, lesion behaviour is inherently heterogeneous. Individual lesions may remain stable, expand slowly or partially regress over time.[10] Whether such heterogeneity persists in SPMS, and how it is distributed within and between patients, has not been systematically examined.

In the present study, we applied a validated longitudinal lesion tracking pipeline to a cohort of patients with SPMS to quantify CLTE over several years of follow-up. [10] We assessed the prevalence and magnitude of chronic lesional activity and its relationship with radiological measures of disease progression. We then investigated whether lesion expansion is associated with microstructural degeneration within lesions[11] and with global markers of neurodegeneration including brain atrophy.[12] Finally, we examined lesion heterogeneity and intra-patient clustering using mixed-effects modelling.

## Methods

### Standard Protocol Approvals and Patient Consents

The study was approved by the University of Sydney Human Research Ethics Committee and conducted in accordance with the tenets of the Declaration of Helsinki. Written informed consent was obtained from all participants.

### Participants

The diagnosis of SPMS was established clinically by the treating neurologist, based on progressive worsening of neurological function sustained over a minimum of 12 months, not attributable to a clinical relapse or new inflammatory lesion activity on MRI. Participants were eligible if they had experienced neither a clinical relapse nor corticosteroid treatment in the 12 months preceding MRI and were on a stable disease-modifying therapy (DMT), or on no therapy, during this period. Demographic and clinical characteristics are summarised in Table 1.

**Table 1.**
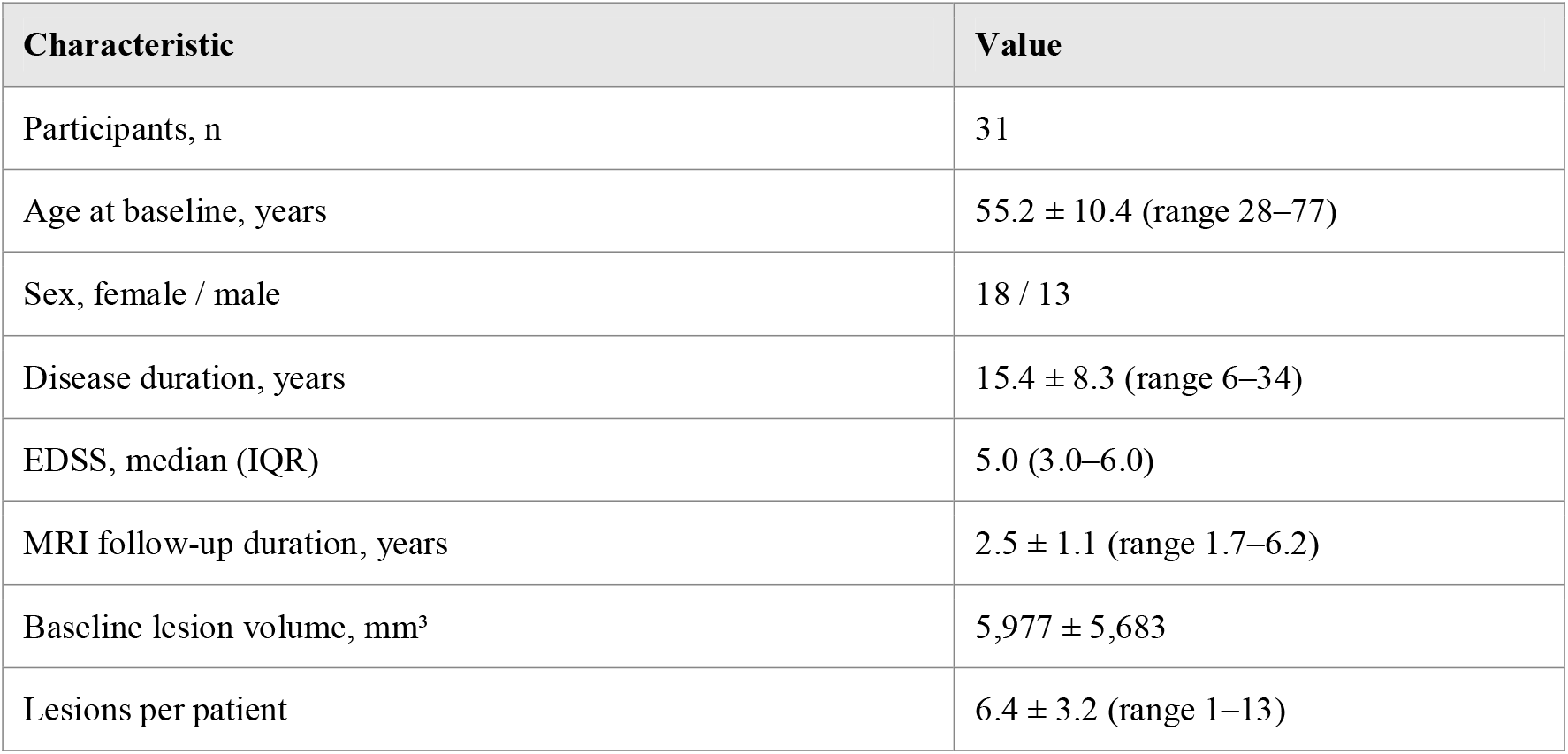
Participant and lesion characteristics. Values are mean ± SD unless stated otherwise. IQR, interquartile range.

| Characteristic | Value |
| --- | --- |
| Participants, n | 31 |
| Age at baseline, years | 55.2 ± 10.4 (range 28–77) |
| Sex, female / male | 18 / 13 |
| Disease duration, years | 15.4 ± 8.3 (range 6–34) |
| EDSS, median (IQR) | 5.0 (3.0–6.0) |
| MRI follow-up duration, years | 2.5 ± 1.1 (range 1.7–6.2) |
| Baseline lesion volume, mm<sup>3</sup> | 5,977 ± 5,683 |
| Lesions per patient | 6.4 ± 3.2 (range 1–13) |

### MRI Acquisition

MRI was performed on a 3T GE Discovery MR750 scanner (GE Medical Systems, Milwaukee, WI) using a standardised longitudinal protocol including pre- and post-contrast 3D T1-weighted imaging, 3D FLAIR, and diffusion-weighted imaging. Detailed acquisition parameters are provided in the Supplementary Material.

### Image Pre-processing

Baseline T1-weighted images were aligned to anterior commissure-posterior commissure (AC-PC) orientation. Follow-up T1, FLAIR, and diffusion tensor images were co-registered to baseline space using FSL and ANTs-based pipelines. [13][14][15] Diffusion MRI data underwent motion, eddy-current, and susceptibility distortion correction prior to tensor reconstruction. Detailed pre-processing procedures are provided in the Supplementary Material.

### Lesion Segmentation

White matter lesions were identified on co-registered T2 FLAIR images and semi-automatically segmented at each time point using JIM 9 software (Xinapse Systems, Essex, UK). All segmentations were performed by a single trained analyst with over 10 years of experience in lesion segmentation (AK) and reviewed for quality assurance.

### Chronic Lesion Tissue Expansion

CLTE was quantified using a validated in-house lesion progression algorithm implemented in Python, as previously described. [10] Briefly, lesions were matched across timepoints based on spatial overlap, while new free-standing and confluent lesions were identified and excluded from CLTE analysis. Follow-up lesion masks were spatially adjusted to account for atrophy-related displacement before lesion volume change was calculated. CLTE was expressed as annualised percentage change (CLTE rate, %/year) and as annualised absolute volumetric change (mm3/year). Patient-level CLTE analyses incorporated all lesions exceeding 50mm^3^ at baseline. New free-standing and confluent lesions detected during follow-up were combined to estimate the patient-level volume of newly formed lesion tissue, enabling direct comparison with expansion-related tissue change.

In a sub-analysis examining lesion behaviour heterogeneity, individual lesions exceeding 100 mm^3^ at baseline were classified according to annualised volumetric change, with the higher volume threshold applied to improve robustness of longitudinal volume estimation. Lesions demonstrating greater than 10% volume increase per year were classified as expanding, those with greater than 10% volume decrease as shrinking, and the remainder as stable.

### Microstructural Analysis

Microstructural tissue damage within chronic lesions was assessed using lesion mean diffusivity (MD) derived from diffusion tensor imaging.[16][11] MD was measured within baseline lesion masks propagated to follow-up space after correction for atrophy-related displacement. Only lesions exceeding 100 mm^3^ at baseline were included. Progressive tissue damage was quantified as the annualised change in MD between time points

### Brain Volumetric Analysis

Ventricular volume was derived from T1-weighted images using AssemblyNet and used as a marker of central brain atrophy (CBA).[17] Global percentage brain volume change was calculated using the SIENA pipeline (Structural Image Evaluation using Normalisation of Atrophy; FSL v4.1; FMRIB, Oxford, UK).[18] T1 intensity inhomogeneity correction was performed, and non-brain tissue was removed using the Brain Extraction Tool (FSL) [19] with manual quality control. Pre-processed brain masks were then processed using SIENA with standard parameters.

### Statistical Analysis

Statistical analyses were performed using Python statistical packages. Continuous variables are reported as mean ± standard deviation (SD) or median with interquartile range (IQR), as appropriate. Longitudinal changes in lesion mean diffusivity were evaluated using the paired t-test. Differences in CLTE rate between sexes were assessed using Welch’s t-test.

Correlations between continuous variables were assessed using Pearson’s correlation coefficient; partial correlation analysis, adjusted for age, sex and disease duration, was used to assess the association between CLTE measures and ventricular volume change as a marker of CBA. Linear regression was additionally used to quantify the magnitude of this relationship. Intra-patient clustering of lesion expansion behaviour was examined using a mixed-effects model with a random intercept for patient, and quantified using the intraclass correlation coefficient (ICC). Statistical significance was set at p < 0.05.

## Results

### Patient Characteristics

Thirty-three patients with SPMS were initially enrolled. Two patients were excluded due to concomitant brain pathology. Data from 31 patients were therefore included in the primary analysis. Participants underwent longitudinal MRI at baseline and follow-up, with a mean follow-up interval of 2.5 ± 1.1 years (range 1.7–6.2 years).

Demographic and clinical characteristics are summarised in Table 1.

Patients were receiving a range of disease-modifying therapies, most commonly ocrelizumab (n = 14), with smaller numbers receiving siponimod, natalizumab, cladribine, ofatumumab, dimethyl fumarate or diroximel fumarate. Four patients were untreated.

### Patient-level analysis of Lesion Expansion

Annualised CLTE values were positive in all patients except one, indicating chronic lesion expansion across nearly the entire cohort. The mean annual lesion expansion rate across the cohort was 7.1 ± 4.1 %. Expansion volume showed variability between patients but was consistently positive, reflecting ongoing enlargement of pre-existing lesions (432 ± 574mm^3^ per year). Exclusion of four cases with small baseline lesion volume did not materially alter the results (6.9 ± 3.9 %; 494 ± 581mm^3^ per year). Figure 1 illustrates the distribution of CLTE rates across individual patients.

**Figure 1.**
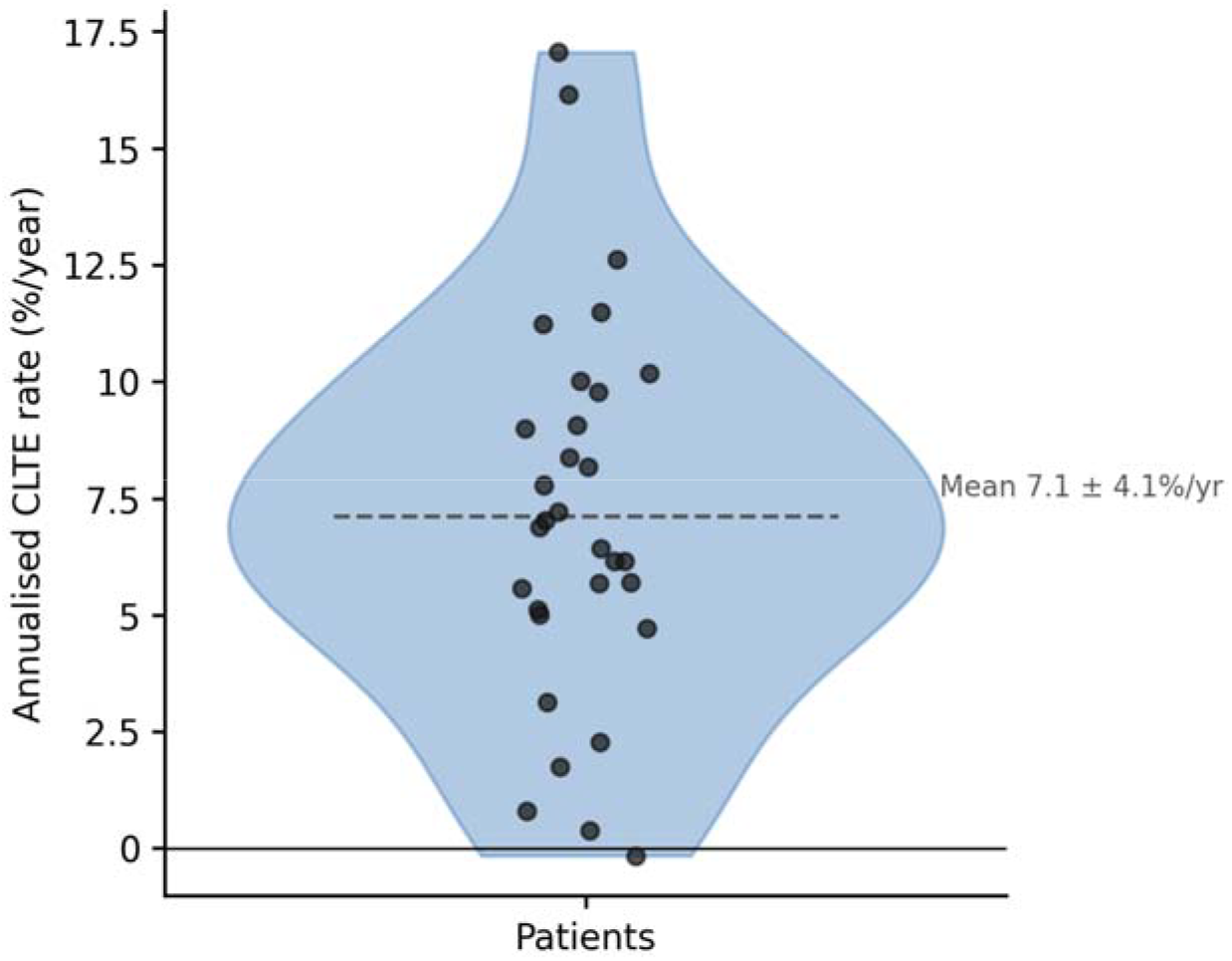
Annualised chronic lesion tissue expansion (CLTE) rates for individual patients, ranked by descending expansion rate. Each dot represents the mean annualised CLTE rate for a patient. The dashed horizontal line indicates the cohort mean (7.1 %/year, SD 4.1).

There was a measurable increase in total lesion volume during the follow-up period, the vast majority of which was attributable to expansion of pre-existing chronic lesions, as illustrated in Figure 2. New lesions, including both free-standing and confluent lesions, were detected in only four patients, with a cumulative volume of 1,501mm^3^. In contrast, chronic lesion expansion accounted for 33,845mm^3^ of tissue increase, representing approximately 96% of the total increase in lesion volume during follow-up.

**Figure 2.**
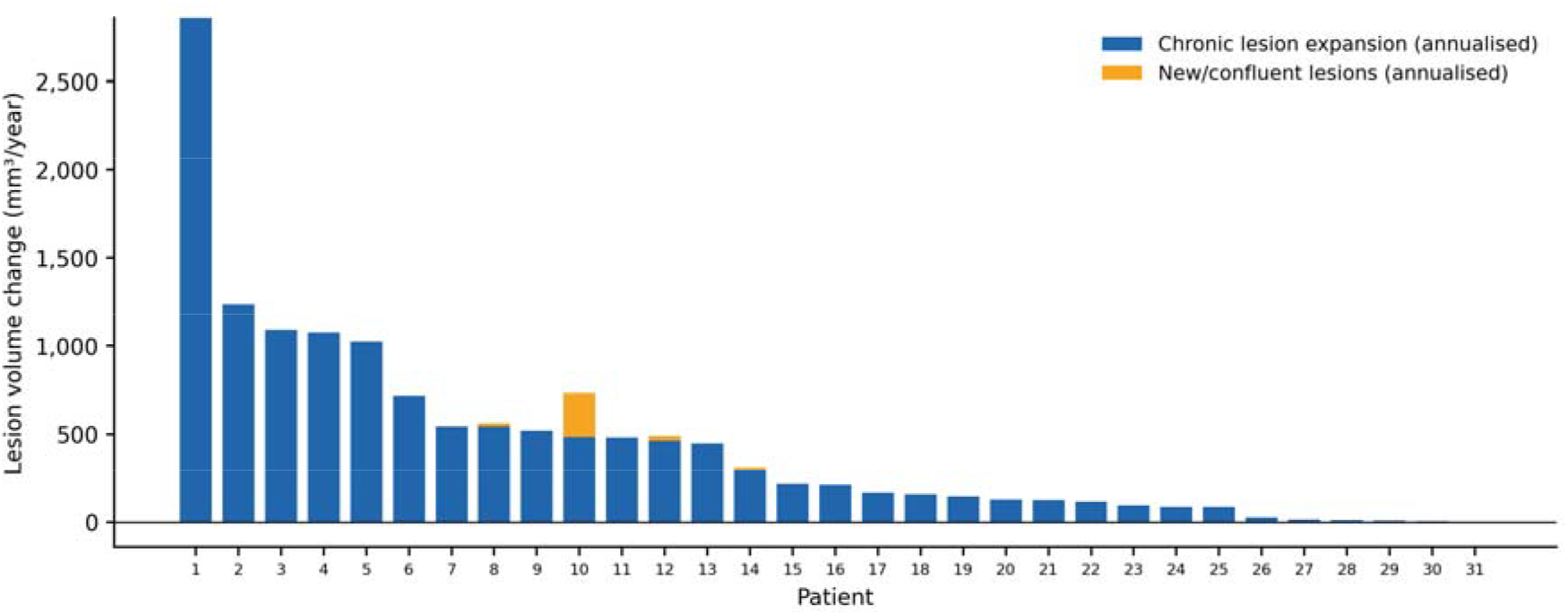
Annual lesion volume change per patient (n = 31), ranked by descending chronic lesion expansion volume. Blue bars: annualised chronic lesion tissue expansion (CLTE) volume (mm^3^/year). Orange bars: annualised volume of newly formed lesions (free-standing and confluent lesions combined). Together these components represent the total annualised increase in lesion-related tissue volume per patient.

This demonstrates that substantial chronic lesion expansion persists in established SPMS despite relatively low levels of new inflammatory lesion formation.

No significant associations were identified between lesion expansion rate and age, sex, disease duration or baseline EDSS (all p > 0.05).

#### Association with Brain Atrophy

CLTE volume showed a significant but moderate association with global brain atrophy measured by SIENA (partial r = −0.36, p = 0.04). In contrast, the relationship with CBA measured by ventricular enlargement was substantially stronger. After adjustment for age, sex and disease duration, annual CLTE volume demonstrated a strong association with ventricular enlargement (partial r = 0.72, p < 0.001).

Linear modelling indicated that 1mm^3^ increase in chronic lesion expansion volume was associated with an approximately 2mm^3^ per year increase in ventricular volume (Figure 3). Exclusion of one participant with extreme expansion burden (residual > 2000 mm^3^) did not materially alter the association (partial r = 0.71, p < 0.001).

**Figure 3.**
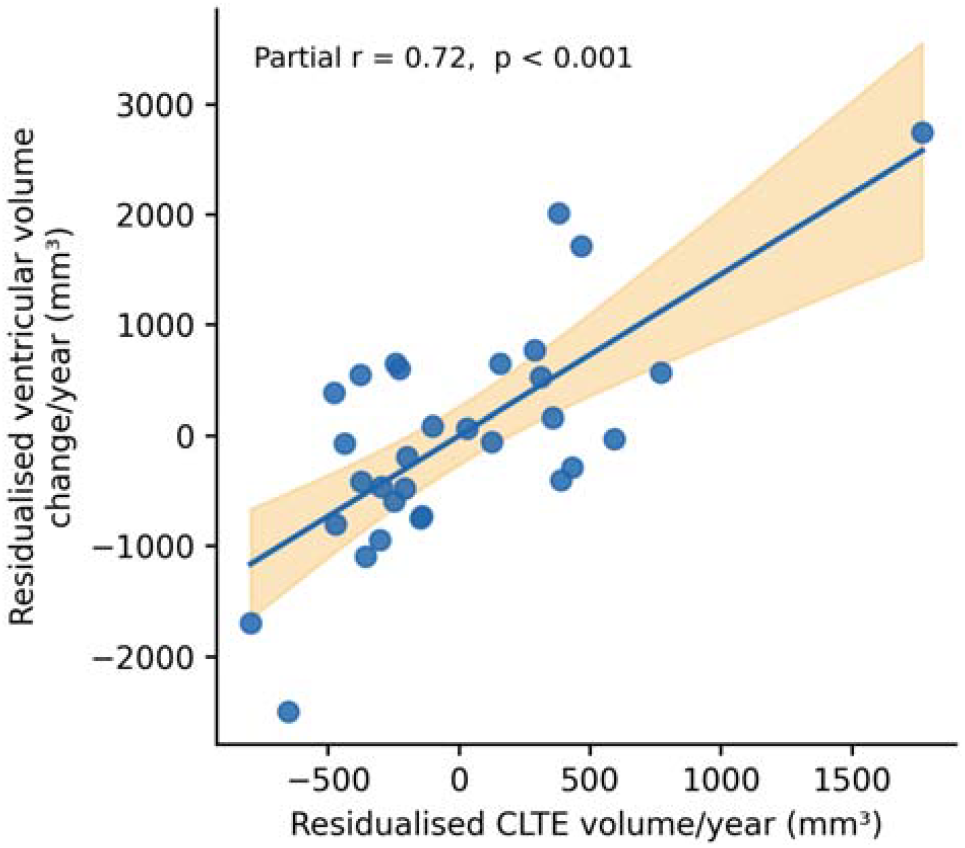
Partial correlation between annual chronic lesion tissue expansion (CLTE) volume and annual ventricular volume change, adjusted for age, sex and disease duration. Each point represents one patient. Both variables are residualised for covariates. The regression line with 95 % confidence interval (shaded) is shown. Partial r = 0.72, p < 0.001.

### Lesion-level Analysis of Lesion Expansion

A total of 186 lesions exceeding 100mm^3^ at baseline were included in the lesion-level analysis (29 patients). The mean number of analysed lesions per patient was 6.4 ± 3.2.

Lesion behaviour was heterogeneous, with 40 (21.5%) of lesions classified as expanding, 145 (78.0%) as stable and 1 (0.5%) as shrinking.

Mixed-effects modelling demonstrated moderate intra-patient clustering of lesion expansion behaviour (ICC = 0.23), indicating that approximately 23% of variance in expansion behaviour was attributable to differences between patients.

### Evolution of Microstructural Change in Chronic Lesions and its Relationship with lesion expansion

One patient was excluded from microstructural analysis due to acquisition-related image corruption, leaving 28 patients and 181 lesions for analysis.

Diffusion MRI demonstrated progressive microstructural change within chronic lesions during follow-up. MD within lesions increased significantly over time (1.222 vs 1.254 ×10□^3^ mm^2^/s, p < 0.001, paired t-test).

In patient-based analysis, the magnitude of MD increase was associated with measures of lesion expansion. Greater increases in MD were observed in patients with higher rates of lesion expansion (r = 0.60, p < 0.001, Figure 4A). A similar association was observed in lesion-based analysis (r = 0.55, p < 0.001, Figure 4B).

**Figure 4.**
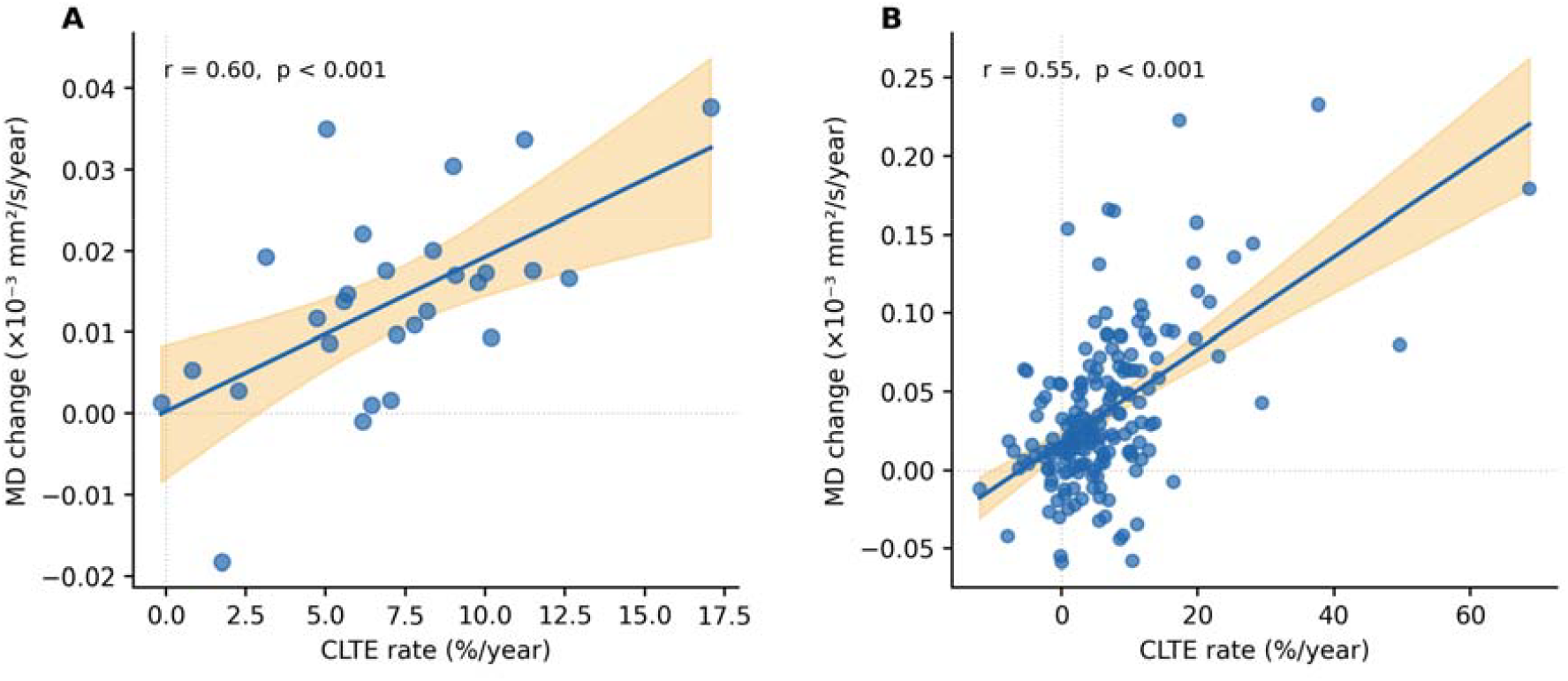
Association between annualised CLTE rate and longitudinal mean diffusivity (MD) change within individual chronic lesions (181 lesions from 28 patients). Left: patient-level analysis; right: lesion-level analysis. Each point represents one patient (left) or one lesion (right). Regression lines with 95 % confidence intervals (shaded) are shown. MD change is expressed as annualised change in MD (×10□^3^ mm^2^/s/year).

## Discussion

This study provides a detailed characterisation of chronic lesion dynamics in secondary progressive multiple sclerosis using longitudinal lesion tracking and microstructural imaging. Several principal findings emerge. First, chronic lesion expansion persists in established SPMS despite minimal new lesion formation. Second, the cumulative tissue change attributable to expansion of existing lesions substantially exceeds the contribution of newly formed lesions. Third, chronic lesion expansion is associated with both microstructural tissue degeneration within lesions and CBA. Together these observations suggest that smouldering inflammatory activity within chronic lesions remains an important contributor to neurodegeneration in progressive multiple sclerosis.

### Persistence of chronic lesion activity in SPMS

The first key observation of this study is that chronic lesion expansion remains prominent in patients with established SPMS. Although overt inflammatory activity decreases during the transition from relapsing to progressive disease, substantial expansion of pre-existing lesions continued to occur over several years of follow up in this cohort.

Importantly, the magnitude of lesion expansion observed in SPMS was similar to that previously reported in relapsing–remitting disease,[7] suggesting that the biological process underlying chronic lesion expansion does not diminish substantially during the transition to the progressive phase of MS.

Complementary evidence for persistent chronic lesion activity in SPMS has also emerged from studies using alternative MRI methods to detect lesion expansion.[8][20] Although these studies used deformation-based approaches rather than direct volumetric measurements, they similarly demonstrated associations between chronic lesion activity and disability progression. Despite methodological differences, the convergence of findings supports the biological relevance of chronic lesion expansion in progressive disease.

These observations align with pathological descriptions of chronic active lesions in progressive multiple sclerosis. Histopathological studies have demonstrated that many long-standing lesions contain a rim of activated microglia and macrophages surrounding a relatively inactive core.[21][22][4] This inflammatory rim is believed to mediate slow outward lesion growth through ongoing demyelination and axonal injury at the lesion edge. The longitudinal imaging findings reported here provide *in vivo* evidence supporting the persistence of this process in advanced disease.

### Chronic lesion expansion as the dominant mechanism of lesion evolution in SPMS

Another central finding was that tissue changes attributable to chronic lesion expansion substantially exceeded the volume of newly formed lesions. Only a small number of patients developed new lesions during follow-up, and their overall volume was minimal compared with tissue change from expansion of pre-existing lesions. These observations indicate that in SPMS the dominant mechanism of lesion-related tissue change is the gradual enlargement of established lesions rather than formation of new inflammatory lesions. Notably, chronic lesion expansion accounted for 96% of lesion burden increase, exceeding previous estimates in relapsing-remitting MS (∼67%).[10] This further emphasises the dominant role of chronic lesion activity in progressive disease.

This finding has important implications for understanding disease progression in SPMS. In relapsing disease, new lesion formation represents the principal mechanism of tissue injury. In contrast, the present results suggest that in progressive disease most ongoing structural damage may arise from slow expansion of existing lesions rather than formation of new lesions.

### Chronic lesion expansion as a driver of tissue destruction and brain atrophy

Microstructural analyses provide further insight into the biological consequences of this process. Mean diffusivity within lesions increased progressively during follow up, and the magnitude of MD change was associated with the rate of lesion expansion. Since MD reflects the mobility of water molecules within tissue and increases when structural barriers such as axons and myelin are lost,[23][11][24][16] the progressive increase in MD observed in expanding lesions is consistent with ongoing tissue rarefaction within the lesion core.

A plausible biological explanation for this imaging pattern is provided by the pathological architecture of chronic active lesions. In these lesions inflammatory activity is concentrated at the lesion rim, where activated microglia and macrophages continue to mediate demyelination and axonal injury. Axonal transection is thought to occur predominantly at the lesion edge, after which the distal intra-lesional segments of damaged axons undergo progressive degeneration within the lesion core. This process leads to gradual loss of structural tissue components and enlargement of the extracellular space.[25][26][27] Because chronic lesions also develop a relatively rigid fibrillary gliotic architecture during the acute stage of lesion formation,[28][29] ongoing tissue loss cannot be compensated for by collapse of the lesion matrix, further contributing to extracellular space expansion.

Importantly, chronic lesion expansion was associated with brain tissue loss and showed a particularly strong relationship with CBA measured by ventricular enlargement. The cumulative volume of lesion expansion demonstrated a robust correlation with ventricular volume increase even after adjustment for demographic and clinical variables. This relationship suggests that chronic lesion expansion may represent an important contributor to central neurodegeneration in progressive multiple sclerosis.

The preferential association with CBA may reflect the predominantly periventricular distribution of chronic expanding lesions.[30][12] Expansion of lesions in these regions is therefore likely to contribute disproportionately to ventricular enlargement.[12] Several mechanisms may underlie the predilection of chronic lesion activity for periventricular regions, including exposure to cerebrospinal fluid-derived inflammatory mediators, reduced remyelinating capacity, hypoxia and compartmentalised microglial activation near the ventricular surface.[31][31][32][33][32][33][33][34] Furthermore, the strong association of lesion expansion with both progressive microstructural degeneration within lesions and CBA suggests that these processes may be mechanistically linked. Inflammatory activity at the lesion rim may lead to axonal transection, with degeneration of intra-lesional axonal segments contributing to tissue rarefaction within the lesion core, reflected by increasing mean diffusivity, while degeneration of extra-lesional periventricular axonal projections within periventricular white matter tracts may contribute to ventricular enlargement through Wallerian and retrograde degeneration.

### Lesion heterogeneity

Marked heterogeneity in lesion behaviour was observed, with most lesions remaining stable or expanding and only rare shrinkage. Mixed-effects modelling demonstrated moderate intra-patient clustering of lesion expansion, indicating that variability was driven predominantly by lesion-specific rather than patient-level factors. These findings support the concept that local lesion microenvironmental factors influence chronic lesion activity and expansion dynamics.

### Implications for disease progression and therapy

Progression in SPMS has traditionally been viewed as predominantly neurodegenerative and largely independent of inflammation due to reduced relapse activity and new lesion formation. However, the present findings suggest that persistent smouldering inflammation within chronic lesions remains a major driver of tissue injury in progressive disease. Chronic lesion expansion in SPMS occurred at rates comparable to relapsing MS and was associated with tissue degeneration and brain atrophy, supporting a model in which inflammatory activity becomes compartmentalised within established lesion rims.

These findings have important therapeutic implications. Current disease-modifying therapies effectively suppress new lesion formation but appear to have limited effects on compartmentalised inflammation within chronic lesions. Given the major contribution of chronic lesion expansion to tissue loss in SPMS, therapies targeting chronic lesion activity and microglial-mediated pathology may be required to slow progression. Quantitative imaging biomarkers of chronic lesion expansion may also provide sensitive outcome measures for such interventions.

### Limitations

Several limitations should be considered. First, the cohort size was modest, although the lesion-level analysis included a large number of individual lesions. Second, lesion segmentation was performed manually, introducing potential operator variability despite careful mask review. Third, diffusion MRI measures provide only indirect markers of tissue integrity and cannot directly demonstrate axonal loss. Fourth, the weaker association with whole-brain atrophy may partly reflect methodological differences between volumetric CLTE measurements and percentage-based SIENA estimates of brain volume loss. Finally, the observational design limits conclusions regarding causal relationships between lesion expansion and brain atrophy. Comparisons of CLTE rates with relapsing-remitting disease are also cross-study in nature, and differences in MRI acquisition and analysis pipelines may limit direct comparability. Furthermore, clinical outcome measures such as disability scores were not included, and the functional consequences of chronic lesion expansion remain to be established.

### Conclusion

Chronic lesion expansion remains a prominent pathological process in SPMS and represents the dominant source of ongoing lesion-related tissue change, substantially exceeding the contribution of new lesion formation. Its association with progressive microstructural degeneration and CBA supports chronic lesion expansion as a key driver of neurodegeneration and disease progression in progressive MS.

## Data Availability

All data produced in the present study are available upon reasonable request to the authors

## Data Availability Statement

Anonymised data is available upon reasonable request.

## Supplementary Material

### MRI Acquisition Parameters and Image Pre-processing

#### S1. MRI Acquisition Parameters

MRI was performed on a 3T GE Discovery MR750 scanner (GE Medical Systems, Milwaukee, WI). The following sequences were acquired at each time point:

a. Pre- and post-contrast (gadolinium) Sagittal 3D T1: GE BRAVO sequence, duration 4 min each, FOV 256 mm, slice thickness 1 mm, TE 2.7 ms, TR 7.2 ms, flip angle 12°, pixel spacing 1 mm. Acquisition matrix (Freq × Phase) 256 × 256, resulting in 1 mm isotropic acquisition voxel size. Reconstruction matrix 256 × 256.
b. FLAIR CUBE; GE CUBE T2 FLAIR sequence, duration 6 min, FOV 240 mm, slice thickness 1.2 mm, acquisition matrix (Freq × Phase) 256 × 244, TE 163 ms, TR 8000 ms, flip angle 90°, pixel spacing 0.47 mm. Reconstruction matrix 512 × 512.
c. Echo-planar imaging based diffusion-weighted MRI, duration 9 min (64 directions with 2 mm isotropic acquisition matrix, TR/TE = 8325/86 ms, b = 1000 s/mm^2^, number of b0 images = 2).

#### S2. Image Pre-processing

##### T1-weighted and FLAIR Images

Baseline T1-weighted images were realigned to anterior commissure–posterior commissure (AC–PC) orientation. Using FLIRT (FSL, FMRIB Software Library, Oxford, UK), [1] follow-up T1 images were co-registered to the baseline AC–PC space by applying transformation matrices derived from linear co-registration between the baseline AC–PC aligned brain and follow-up native T1 brain images. FLAIR images at each time point were then linearly co-registered to the corresponding T1 AC–PC images.

##### Diffusion MRI

Diffusion MRI data were corrected for motion and eddy-current distortion using FSL. Susceptibility-related EPI distortion was minimised by applying deformation maps generated from nonlinear co-registration of the diffusion-weighted b0 image to the T1-weighted image at each time point, using ANTs (Advanced Normalization Tools).

[2] Tensor reconstruction was subsequently performed using MRtrix3. [3] Diffusion tensor images were then linearly co-registered to the corresponding T1 AC–PC space.

FSL, FMRIB Software Library; ANTs, Advanced Normalization Tools; AC–PC, anterior commissure–posterior commissure; FOV, field of view; TE, echo time; TR, repetition time; DWI, diffusion-weighted imaging.

